# Using diffusion MRI to relate hippocampal subfield microstructure to delayed verbal memory in cognitively intact individuals at genetic risk for developing Alzheimer’s disease

**DOI:** 10.1101/2025.04.07.25325398

**Authors:** Jennapher Lingo VanGilder, Andrew Hooyman, Sasha Hakhu, Kurt G. Schilling, Leland S. Hu, Yuxiang Zhou, Richard J. Caselli, Leslie C. Baxter, Scott C. Beeman

**Affiliations:** School of Biological and Health Systems Engineering, Arizona State University; Department of Physical Therapy, Chapman University, Irvine, CA; Vanderbilt University Institute of Imaging Science, Vanderbilt University, Nashville, TN; Department of Radiology, Mayo Clinic, Phoenix, AZ; Department of Medical Physics, Mayo Clinic, Phoenix, AZ; Department of Neurology, Mayo Clinic, Phoenix, AZ; Department of Psychiatry & Psychology, Mayo Clinic, Phoenix, AZ

## Abstract

Early intervention to delay the onset of Alzheimer’s disease (AD) is an important treatment strategy but detecting at-risk individuals before significant disease progression remains challenging. This study evaluates the relationship between hippocampal microstructure and verbal cognition in cognitively intact older adults, focusing on the differences between APOΕ ε4 allele carriers and noncarriers. Participants (n=41 noncarriers, 33 carriers) over 60 years old (mean±SD: carriers 71±6.6; noncarriers 71±6.4 years) underwent diffusion-weighted magnetic resonance imaging (dMRI) and volumetric assessments. We assessed hippocampal structure, including microstructure, using Neurite Orientation Dispersion and Density Imaging (NODDI), diffusion tensor imaging (DTI), and volumetric measures. Regression analyses examined the relationship between these hippocampal measures and verbal and visuospatial cognition, as evaluated by the Rey delayed recall tests i.e., the Auditory Verbal Learning Test (AVLT) and Complex Figure Test (CFT), respectively. Results indicated that while volumetric data showed no significant findings, microstructural measures, particularly orientation dispersion (ODI) in the left subiculum, were positively associated with verbal recall in APOΕ ε4 carriers (FDR-corrected p=0.01). These findings suggest that hippocampal microstructure, rather than volume, may provide early insights into cognitive decline in individuals at genetic risk for AD.

## Introduction

While there is currently no cure for Alzheimer’s disease (AD), treatment can be administered to delay its progression and onset of symptoms (Nelson and Tabet, 2015). A major challenge of this treatment strategy has been our inability to accurately detect at-risk individuals prior to significant disease progression. The apolipoprotein epsilon 4 (APOE ε4) allele is a common genetic risk factor for developing AD and is thought to be associated with synaptic toxicity and eventual neuronal loss (Hesse et al., 2019; Lane-Donovan and Herz, 2017) through beta amyloid protein accumulation (Koffie et al., 2012; Sheng et al., 2012). Current hypotheses of AD pathogenesis posit that the hippocampus becomes functionally disconnected from neural networks due to synaptic loss (Kashyap et al., 2019; Li and Selkoe, 2020), pathology begins within left hemispheric structures (e.g., the hippocampal complex (Abraham et al., 2020; Jessen et al., 2006; Mohtasib et al., 2022)), and that delayed verbal memory is one of the first cognitive functions to demonstrate impairment in APOE ε4 carriers (Caselli et al., 2009).

In line with these hypotheses, we have recently shown that reduced functional connectivity of the left (but not right) hippocampus was significantly related to decline in delayed verbal memory performance in cognitively intact APOE ε4 carriers; notably, this effect was independent of hippocampal volumes, nonverbal memory change, and was not observed in noncarriers (Baxter et al., 2023). These findings suggest that APOE ε4 carriers may demonstrate pathological changes in left hippocampal functional connectivity earlier than changes in hippocampal volumes and prior to frank behavioral impairment. Given that APOE ε4 carriers may experience exacerbated synapse loss ahead of aging, it is plausible that the functional hippocampal disconnection may be due to a loss of synaptic connections, although the microstructural correlates of this brain-behavioral relationship have not been well-described in intact subjects.

Radiological interrogation of the hippocampus as a singular neural region may be insensitive for detection of subtle brain-behavioral relationships, given that subregions (or subfields) within the hippocampus are histologically (Adler et al., 2014; Flores et al., 2020) and functionally (Chang et al., 2021; Zammit et al., 2017) distinct, and that hippocampal subfields (i.e., the cornu ammonis (CA) 1, CA2-3, CA4-dentate gyrus, and the subiculum) may be differentially impacted by AD-related pathology. Reports from volumetric studies have linked subicular atrophy to declines in delayed verbal memory (Lindberg et al., 2017), as well as to conversion from mild cognitive impairment to AD (Abraham et al., 2020; Carlesimo et al., 2015; Hett et al., 2019; Zhao et al., 2019), whereas other reports suggest that CA1 atrophy may be the best indicator of conversion to AD (Apostolova et al., 2006; de Flores et al., 2015). While collectively these studies implicate the role of hippocampal subfield volumes in predicting AD progression in clinical populations, degeneration of hippocampal microstructure may begin years prior to our ability to measure it with standard clinical neuroimaging methodologies. The relationship between declines in memory and subfield *microstructure* in APOE ε4 carriers is less studied and may confer greater sensitivity to subtle early changes.

Quantitative multi-shell diffusion magnetic resonance imaging (dMRI) can be made exquisitely sensitive to the underlying microstructure of dendrites and axons (i.e., components of the synapse) through rational sampling of the dMRI parameter space and biophysical modeling of diffusion properties within brain tissues. Diffusion Tensor Imaging (DTI) and Neurite Orientation Dispersion and Density Imaging (NODDI) are two methods to assess white matter anatomy. DTI is the clinical standard-of-care model used to calculate diffusion metrics such as fractional anisotropy (FA, a measure of white matter integrity), and radial diffusivity (RD, a measure of diffusion especially in the radial direction of a white matter tract), although it lacks specificity; for instance, various microstructural alterations such as demyelination or axonal loss can impact the RD metric (Jelescu et al., 2020) such that the underlying physiological mechanism is unclear. Advanced diffusion models like NODDI have been developed to address these limitations, offering additional metrics like neurite density index (NDI) and orientation dispersion index (ODI) (Qiu et al., 2017). NDI offers a more direct measure of the density of neurites (axons and dendrites), providing insight into neuronal integrity. ODI quantifies the degree of dispersion of axon orientations, helping to capture changes in tissue microstructure that may not be detected by traditional models, particularly in areas with complex fiber architecture. Importantly, measures of ODI correlate with PET-based measures of synaptic density (Kohli et al., 2021; Mak et al., 2021) and both NODDI metrics have been shown to distinguish individuals with mild cognitive impairment from those with AD (Fu et al., 2020; Vogt et al., 2020), but whether it can be used to identify microstructural degeneration in asymptomatic individuals *at-risk* for developing AD is unknown.

The purpose of this study was to extend our previous work (Baxter et al., 2023) that showed left hippocampal connectivity was associated with changes in memory performance in APOE ε4 carriers. Here, we systematically evaluated if structural measures of specific hippocampal subfields were associated with memory in this cognitively intact but at-risk group. In alignment with our prior findings, we hypothesized that reduced left hippocampal microstructure would predict poorer delayed verbal memory scores in APOE ε4 carriers.

Given that volumetric studies suggest the subiculum is first to show structural changes (Chi et al., 2022; Zammit et al., 2017), we hypothesized that NODDI-based measures the left subiculum microstructure would preferentially predict memory performance in cognitively intact APOE ε4 carriers.

## Methods

All experimental procedures were approved by the Institution’s Review Board and adhered to the Declaration of Helsinki. Participants provided informed consent prior study enrollment. All data were collected as part of the longitudinal AZ APOE study by the Mayo Clinic in Arizona; allelic status was determined upon enrollment (see Caselli et al., 2011, 2010)). Neuroimaging and cognitive data from 74 community-dwelling adults (carriers n=41; noncarriers n=33) with mean ± SD age of 71±6.6 and 71±6.4 years, respectively, were retrospectively analyzed for this follow-up study. Cognitively intact status was determined by performance on the MMSE (Folstein et al., 1975), Instrumental Activities of Daily Living Questionnaire (Katz et al., 1970), and Hamilton Depression Rating Scale (Hamilton, 1967). A licensed neuroradiologist, neurologist, and neuropsychologist reviewed the data to screen for the presence of confounding medical, neurological, and/or psychiatric disorders and conversion to mild cognitive impairment. To minimize any age-related structural effects, we included those participants aged 60 year and above. The delayed recall measure from the Rey Auditory Verbal Learning Test (AVLT-DR) was used due to its sensitivity in other studies of preclinical AD (Caselli et al., 2009). Since there is a left-right bias for verbal-visual memory, respectively, we also examined similar effects for Rey-Osterrieth Complex figure test delayed recall scores (CFT-DR). Cognitive and imaging data were collected within a few weeks of each other.

### MRI acquisition and preprocessing

Participants underwent multi-shell diffusion imaging on a Siemens 3T MRI Skyra scanner at Mayo Clinic, Scottsdale, AZ. An EPI sequence was used to acquire diffusion-weighted images with the following parameters: b-values = 0.5, 1, and 2 ms/µm^2^ with 6, 38, and 47 diffusion encoding directions, respectively, and nine interleaved b=0 volumes; TR/TE: 4.3/0.101 s; flip-angle = 90°; 2x2x2 mm^3^ spatial resolution; slice thickness: 2.0 mm; scan time 7.87 minutes). T1-weighted images were also acquired (parameters: TR/TE: 6.7/3.1 s, flip-angle=9°, FOV=256 mm, 1x1x1 mm^3^ spatial resolution, 208 sagittal slices) and used to nonlinearly align images during the coregistration process and for volumetry. FSL (FMRIB, Oxford, UK) and was used to reduce noise (dwidenoise), and susceptibility-induced distortion (Schilling et al., 2020), to create a whole brain mask to extract brain from non-brain tissues (dwi2mask) and to correct for motion and eddy currents (eddy). To account for the rotational component of registration, the b-vector files were compensated after motion correction and prior to calculating the b matrices. B1-field inhomogeneity across all diffusion volumes was then corrected for (dwibiascorrect) in MRtrix (Tournier et al., 2019).

Neurite Orientation Dispersion and Density Imaging (NODDI) is a well-established model of dMRI that quantifies the net density (neurite density index, NDI) and orientation (orientation dispersion index, ODI) of dendrites and axons within each voxel (Zhang et al., 2012). The NODDI model (Zhang et al., 2012) was applied to each acquisition to quantify the orientation dispersion and neurite density indices in each voxel using MATLAB (MathWorks, Inc.). ODI is a value that ranges from 0 to 1 and is interpreted as a measure of neurite coherence, wherein a value closer to 1 suggests that the net orientation of neurites within that voxel is widely dispersed and a value closer to 0 suggests that the net orientation has coherence in a uniform direction; NDI is the volume fraction of tissue that comprises neurites, or neurite density, where a value closer to 1 would suggest that the intravoxel tissue is comprised largely of neurites. We also calculated fractional anisotropy (FA) and mean diffusivity (MD) maps using the standard of care DTI model (FSL’s DTIFIT tool).

Each participant’s b0 image was nonlinearly transformed to their structural T1-weighted image (FSL’s fnirt), which was then nonlinearly transformed to the Montreal Neurological Institute 152 (MNI) standard brain atlas. Bilateral hippocampi (Harvard-Oxford cortical and subcortical structural atlases (Desikan et al., 2006; Frazier et al., 2005; Makris et al., 2006)) and subfields (i.e., the cornu ammonis (CA) 1, CA 2-3, CA 4-dentate gyrus, and the subiculum (Iglesias et al., 2015)) were included as hippocampal regions of interest (ROIs). To account for age-related differences in brain structure such as ventricular expansion (Gordon et al., 2013), the ROIs were nonlinearly transformed to an older adult brain atlas (described in (Gordon et al., 2015)) and then to participant native diffusion space. To constrain our measures to only gray matter voxels, ROIs were thresholded by each participant’s grey matter tissue mask (FAST). Volume (fslstats) and NODDI indices were then quantified for each ROI and the tissue-weighted mean was calculated to reduce estimation bias caused by partial volume effects (Parker et al., 2021). To address cerebrospinal fluid (CSF) contamination in our ROI analysis, we implemented a tissue-weighted mean approach (Parker et al., 2021). Traditional methods calculate the mean NODDI metric by averaging all voxels in an ROI, which can be biased by CSF contamination. Our method uses tissue fraction metrics from NODDI to account for varying CSF contamination across voxels, providing a more accurate representation of regional microstructure. This approach corrects for CSF partial volume effects while maintaining the intent of estimating the average microstructure within the tissue of the ROI.

Building on earlier findings that memory trajectories for APOE-ε4 heterozygote carriers start to diverge from noncarriers around age 60 (Baxter et al., 2023), and to minimize significant age-related brain differences when combining younger and older adults, we constrained our analyses to participants aged 60 and above.

### Statistical analysis

R Version 2024.04.2+764 was used for statistical analyses. To evaluate the extent to which APOE ε4 status predicts the relationship between hippocampal regions and delayed verbal memory, we conducted a series of iterative linear regression models. These models included participant allelic group (APOE ε4 carrier/noncarrier), diffusion imaging metrics (for example, ODI or FA) for each region of interest (ROI), and their interaction as predictor variables for CFT-DR and AVLT-DR scores. Participant age was included as a covariate to account for age-related differences in tissue microstructure. Separate regressions were performed for imaging metrics across each ROI. To correct for multiple comparisons, a false discovery rate (FDR) correction was applied. For any significant hippocampal region-by-APOE ε4 status interaction, a follow-up sensitivity analysis was conducted to assess whether the same relationship persisted when considering the volumetric measure of the hippocampal region. This analysis aimed to determine whether the microstructural metrics (ODI or NDI) were better at detecting the influence of APOE ε4 status on cognitive performance compared to traditional volumetric measures. Additionally, p-values from the APOE interaction were log-transformed and plotted across the 10 hippocampal ROIs for FA, ODI, and volume metrics in relation to CFT recall and AVLT scores, with Bonferroni threshold for 10 tests (alpha = .005) applied to assess significance.

## Results

In our analysis of the APOE interaction across 10 hippocampal regions of interest, including left and right CA1, CA2-3, CA4, subiculum, and the whole hippocampus, we assessed FA, ODI, and volumetric metrics in relation to delayed recall CFT-DR and AVLT-DR scores). Demographic and NODDI data are summarized in Table 1.

**Table 1.**
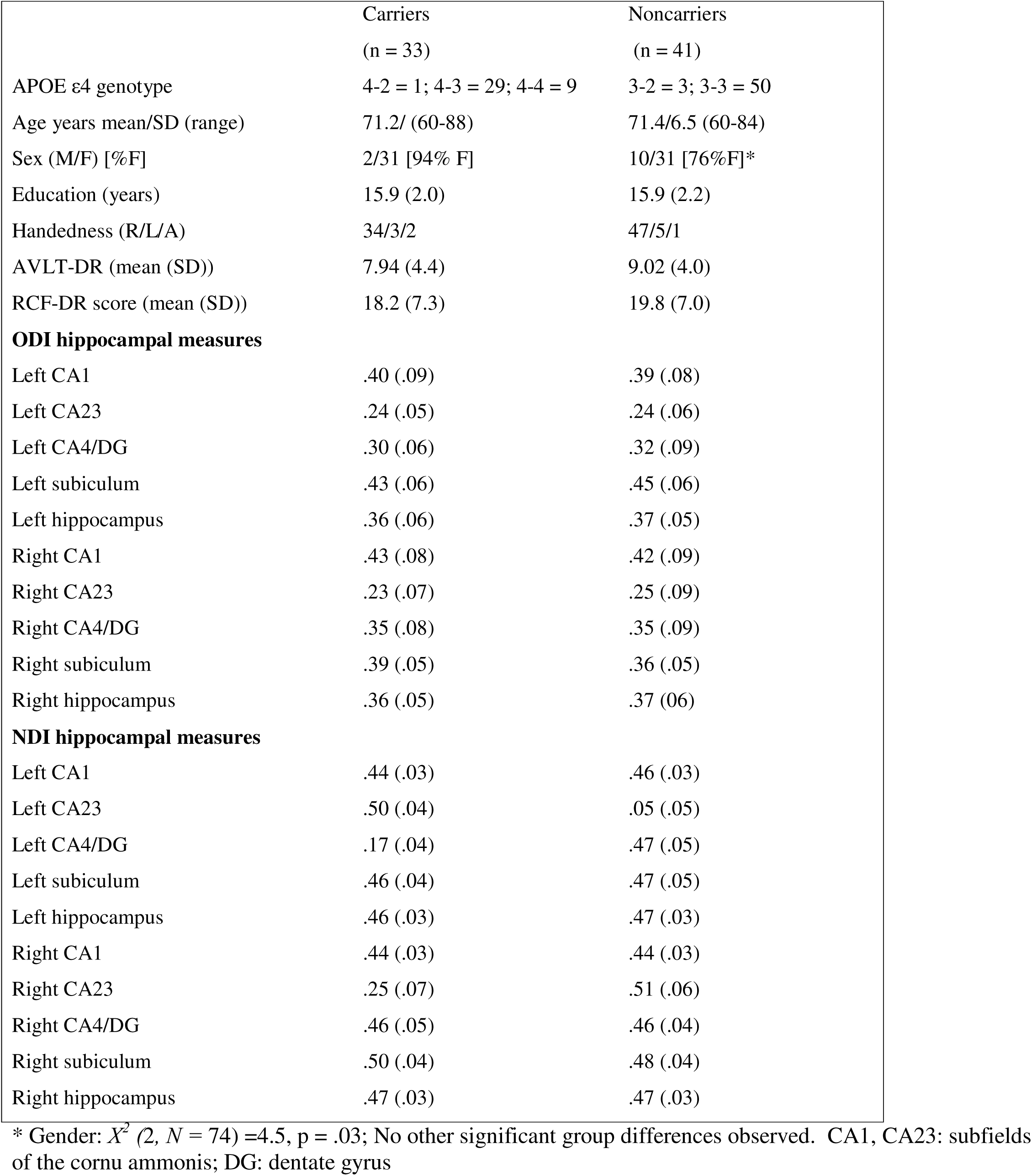
Demographic and NODDI data.

|  | Carriers<br>(n = 33) | Noncarriers<br>(n = 41) |
| --- | --- | --- |
| APOE ε4 genotype | 4-2 = 1; 4-3 = 29; 4-4 = 9 | 3-2 = 3; 3-3 = 50 |
| Age years mean/SD (range) | 71.2/ (60-88) | 71.4/6.5 (60-84) |
| Sex (M/F) [%F] | 2/31 [94% F] | 10/31 [76%F]* |
| Education (years) | 15.9 (2.0) | 15.9 (2.2) |
| Handedness (R/L/A) | 34/3/2 | 47/5/1 |
| AVLT-DR (mean (SD)) | 7.94 (4.4) | 9.02 (4.0) |
| RCF-DR score (mean (SD)) | 18.2 (7.3) | 19.8 (7.0) |
| **ODI hippocampal measures** |  |  |
| Left CA1 | .40 (.09) | .39 (.08) |
| Left CA23 | .24 (.05) | .24 (.06) |
| Left CA4/DG | .30 (.06) | .32 (.09) |
| Left subiculum | .43 (.06) | .45 (.06) |
| Left hippocampus | .36 (.06) | .37 (.05) |
| Right CA1 | .43 (.08) | .42 (.09) |
| Right CA23 | .23 (.07) | .25 (.09) |
| Right CA4/DG | .35 (.08) | .35 (.09) |
| Right subiculum | .39 (.05) | .36 (.05) |
| Right hippocampus | .36 (.05) | .37 (.06) |
| **NDI hippocampal measures** |  |  |
| Left CA1 | .44 (.03) | .46 (.03) |
| Left CA23 | .50 (.04) | .05 (.05) |
| Left CA4/DG | .17 (.04) | .47 (.05) |
| Left subiculum | .46 (.04) | .47 (.05) |
| Left hippocampus | .46 (.03) | .47 (.03) |
| Right CA1 | .44 (.03) | .44 (.03) |
| Right CA23 | .25 (.07) | .51 (.06) |
| Right CA4/DG | .46 (.05) | .46 (.04) |
| Right subiculum | .50 (.04) | .48 (.04) |
| Right hippocampus | .47 (.03) | .47 (.03) |

\* Gender: $X^2(2, N = 74) = 4.5$ , $p = .03$ ; No other significant group differences observed. CA1, CA23: subfields of the cornu ammonis; DG: dentate gyrus

Results from the iterative modeling demonstrated that only the left subiculum exhibited a statistically significant interaction with AVLT scores after Bonferroni correction (p<0.005) (Figure 1). Further comparison between APOE ε4 carriers and noncarriers showed a significant association between delayed recall scores and left subiculum ODI (z-score) in carriers, a relationship not observed in noncarriers (Figure 2). Statistical results of these relationships between imaging metrics and cognitive scores based on carrier status are summarized in Tables 2 and 3. Supplementary analyses demonstrated the relationship between DTI-FA and NODDI-Kappa (a component measure of neurite orientation dispersion) within the left subiculum (Figure S1).

**Figure 1.**
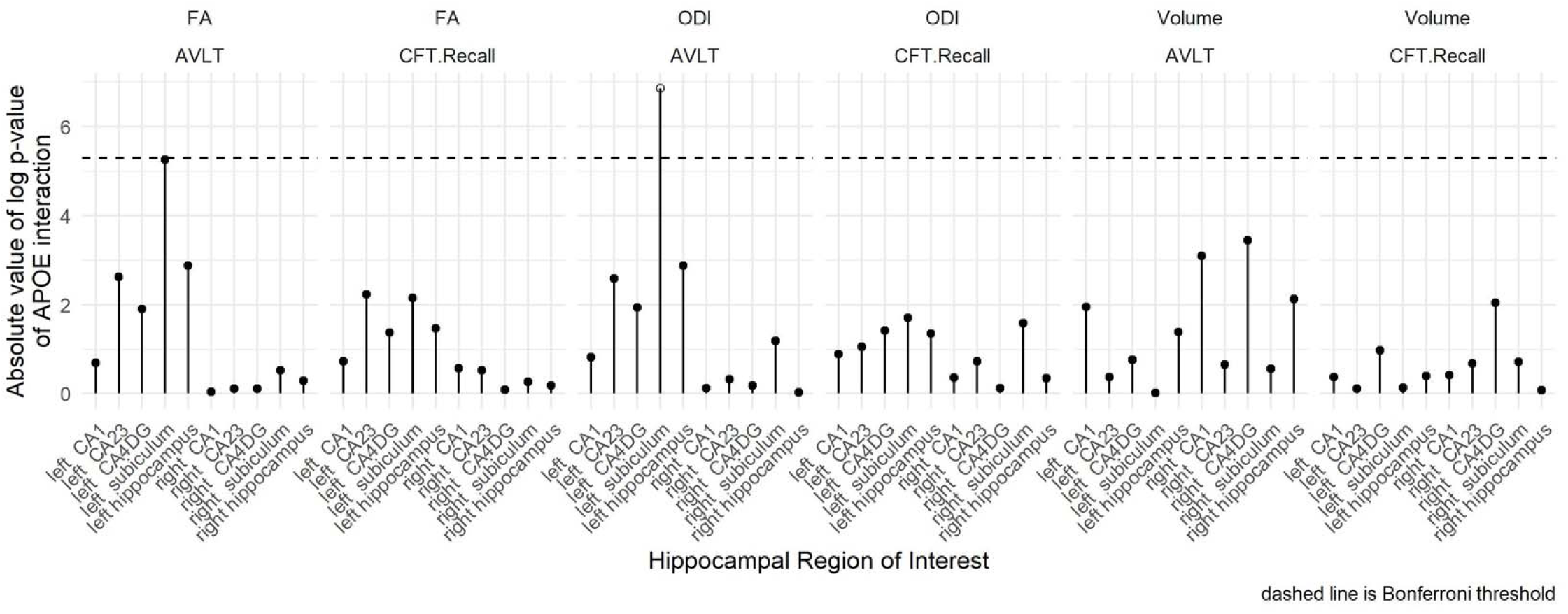
Shown are the absolute values of log-transformed raw p-values for the APOE ε4 interaction across 10 hippocampal regions of interest (i.e., left and right CA1, CA2-3, CA4, subiculum and whole hippocampus), assessed for FA, ODI, and volume metrics in relation to CFT recall and AVLT scores. Higher the magnitudes on the graph correspond to smaller p-values. The dashed line represents the threshold for statistical significance after Bonferroni correction for 10 comparisons (p=0.005). Notably, only the left subiculum was associated with AVLT, indicating significant interaction effects that persist beyond multiple comparison correction.

**Figure 2.**
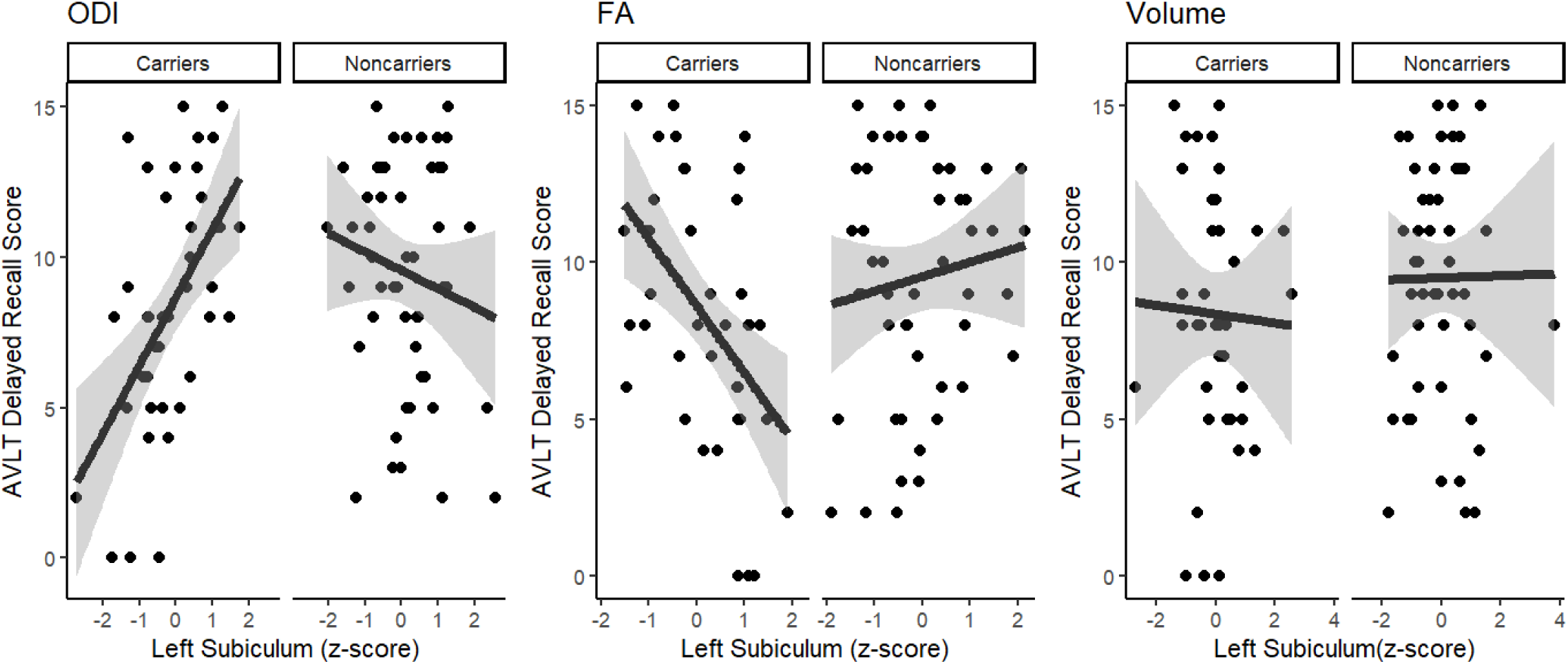
Shown are comparisons of AVLT scores to microstructure imaging metrics (NODDI-ODI and DTI-FA) and volume measures for the left subiculum region amongst APOE ε4 carrier and non-carrier groups. Cognitively intact APOE ε4 carriers demonstrated a significant relationship between Delayed Recall scores and left subiculum ODI (z-score), whereas noncarriers did not.

**Table 2.** Summary based on statistical relationship (multiple comparisons corrected p-values) between imaging metrics (ODI, FA and Volume) and cognitive test (AVLT and CFT).

| Region | ODI &AVLT | ODI & CFT | FA & AVLT | FA & CFT | Volume & AVLT | Volume & CFT |
| --- | --- | --- | --- | --- | --- | --- |
| Left CA1 | 0.4422 | 0.4124 | 0.5031 | 0.5885 | 0.143 | 0.694 |
| Left CA2-3 | 0.0751 | 0.3465 | 0.0732 | 0.7168 | 0.6891 | 0.8925 |
| Left CA4 | 0.1434 | 0.2407 | 0.1489 | 0.8012 | 0.4682 | 0.3778 |
| Left Subiculum | **0.0011\*** | 0.1827 | 0.0052 | 0.9779 | 0.9842 | 0.8756 |
| Left Hippocampus | 0.0563 | 0.2581 | 0.0562 | 0.8223 | 0.2507 | 0.6768 |
| Right CA1 | 0.889 | 0.7 | 0.9639 | 0.616 | 0.0454 | 0.6568 |
| Right CA2-3 | 0.7224 | 0.4853 | 0.892 | 0.6946 | 0.5224 | 0.5065 |
| Right CA4 | 0.8312 | 0.8837 | 0.8909 | 0.623 | 0.032 | 0.1299 |
| Right Subiculum | 0.3073 | 0.2042 | 0.5942 | 0.8696 | 0.5728 | 0.492 |
| Right Hippocampus | 0.9749 | 0.7052 | 0.7458 | 0.6685 | 0.1193 | 0.9285 |

\*p-values<0.05 are considered statistically significant

**Table 3.** Summary of statistical relationships ((multiple comparisons corrected p-values and R^2^ values) between imaging metrics (ODI, FA, and Volume) and cognitive performance (AVLT) in APOE-Ε4 carrier and non-carrier groups.

| METRIC | APOE-E4<br>CARRIER<br>STTAUS | R<sup>2</sup> | p-value |
| --- | --- | --- | --- |
| FA | Non-carrier | 0.0158 | 0.3842 |
| FA | Carrier | 0.2243 | 0.0018 |
| ODI | Non-carrier | 0.0256 | 0.2673 |
| ODI | Carrier | 0.2937 | 0.0003 |
| Vol | Non-carrier | 0.0001 | 0.9501 |
| Vol | Carrier | 0.002 | 0.7837 |

## Discussion

This study found that a measure of hippocampal microstructure significantly associated with verbal memory performance in only older adults who carry the APOE ε4 allele. The association of microstructural integrity and verbal memory was for the left but not right hippocampus, and as expected, there was no association of microstructural measures with visual memory. These findings point to early microstructural changes in cognitively intact individuals with genetic risk for AD that are in alignment with our previous findings indicating weak left hippocampal functional connectivity (Baxter et al., 2023). Thus, using these advance imaging techniques, both structural and functional evidence of hippocampal associations with memory functioning in intact at-risk older adults can be observed prior to cognitive decline in the absence of hippocampal volumetric associations. Indeed, we hypothesize that these microstructural associations with memory function precede eventual volumetric associations; this will be the subject of future investigation. The expected association of age and memory performance was observed for both carriers and non-carriers. Since the microstructural-memory association was not found for the noncarriers, this finding more likely represents early pathological changes in the at-risk group.

Hippocampal subfield segmentation indicated that subiculum microstructure was most strongly correlated with verbal memory performance. Functional MRI studies have observed greater activation within the subiculum during memory retrieval (Gabrieli et al., 1997; Ledergerber et al., 2021) and volumetric studies implicate subicular atrophy with delayed memory performance in older adults (Chi et al., 2022; Zammit et al., 2017) and individuals at-risk for developing AD (Carlesimo et al., 2015). Recent volumetric studies found reduced left subiculum volumes associated with poorer performance on the DR in APOE ε4 carriers who had converted to mild cognitive impairment (Abraham et al., 2020) and in individuals with subjective cognitive decline (Zhao et al., 2019) and AD (Sarica et al., 2018).

In its simplest form, diffusion weighted imaging can be made sensitive to the *apparent* diffusivity of water within tissue, quantified as the apparent diffusion coefficient (ADC) which reflects the true self-diffusion coefficient of water weighted by the boundary-driven restriction of water displacement within a voxel.

Importantly, in complex biologic tissue like brain, where microstructural boundaries like myelin sheaths can preferentially restrict water displacement in one direction (radial) and permit displacement in another (axial), diffusion can be restricted as a function of direction. Thus, when collected at various angles relative to the tissue, apparent diffusivity can vary as a function of tissue microstructure and angle of diffusion weighting. Typically, this phenomenon is quantified by the collection of multiple diffusion-weighted volumes, each with a diffusion weighting vector pointed in a different direction in space, and the calculation of a tensor to determine the mean diffusivity, degree of anisotropy, and principal direction of diffusion within individual voxels. This so-called diffusion tensor imaging method is ubiquitous amongst academic and clinical research and has long been employed in the study of AD pathogenesis (Chen et al., 2023; Cheng et al., 2023; Lancaster et al., 2016; Maggipinto et al., 2017; Zhu et al., 2015) . However, the sensitivity of DTI to early AD-associated microstructural deficits and the specificity of DTI to precise cell structure differences are lacking; both issues stemming from the very nature of DTI, which is merely a quantitative representation of the *diffusion signal*, not a biophysical model design to quantify tangible microstructure (e.g., neurite density and orientation).

Our findings highlight that hippocampal microstructure, rather than volume, is associated with AVLT- DR memory scores in APOE ε4 carriers. This suggests early microstructural alterations in at-risk individuals, but the specific biological mechanisms underlying these associations remain unclear. Contrary to our observed association between ODI and memory performance, we did not observe a significant association between NDI and memory performance. Furthermore, we did not observe significant group differences in ODI, NDI, or MD between carriers and non-carriers. NDI is commonly interpreted as a marker of neurite density and tissue integrity, so the lack of group differences raises the question of why NDI does not appear to be a key factor in verbal memory. This suggests that other microstructural properties—such as myelination—may play a more prominent role. Prior research (Fukutomi et al., 2019) has shown that NDI is inversely related to MD, a measure often linked to overall microstructural degradation (Vernooij et al., 2009). However, in the absence of group differences in these metrics, our findings suggest that hippocampal microstructural differences relevant to memory may not be driven by neurite density alone, but rather by factors influencing neurite organization and white matter integrity. This aligns with evidence that hippocampal connectivity and fiber structure, rather than solely neuronal packing density, are critical for memory function (Nazeri et al., 2015).

Additionally, our results indicate a relationship between verbal memory and ODI in APOE ε4 carriers, but no significant group difference in ODI values. This raises the question of whether it is appropriate to describe this association as “pathological.” Since ODI values in carriers were comparable to noncarriers, this suggests that observed differences may reflect individual variation in hippocampal microstructure rather than overt disease-related degeneration. Moreover, prior research underscores the importance of hippocampal integrity for verbal memory not only in at-risk individuals but also in cognitively normal adults (Qian et al., 2020). Thus, the observed association may reflect a broader relationship between hippocampal organization and memory, rather than early pathological changes. Future longitudinal studies could investigate whether the observed relationship between hippocampal ODI and memory reflects early compensatory mechanisms or is independent of neurodegenerative processes by examining its association with established markers of pathology and longitudinal cognitive decline.

To better interpret the role of ODI in this context, we examined its relationship with FA, as both metrics provide complementary insights into hippocampal microstructure. While FA captures overall diffusion directionality, ODI and Kappa provide more detailed information about neurite complexity and fiber coherence. Given that ODI was linked to memory performance, understanding its relationship with FA may help clarify whether microstructural factors beyond traditional diffusion metrics contribute to cognitive function. Our findings suggest that while traditional DTI metrics capture basic diffusion properties, incorporating advanced models like NODDI may provide deeper insights into microstructural contributions to cognition. Future studies could explore how combining these measures improves predictions of cognitive outcomes.

Rational design of dMRI experiments and biophysical modelling of restricted diffusion within tissues (and the expected dMRI signal in such a modelled system), allows for non-invasive quantification of specific microstructure. In this case, we employ the already well-established NODDI model to investigate the differences in neurite microstructure associated with APOE ε4 allele carriership in cognitively intact research participants. Previous efforts have shown that NODDI measures correlated with tau pathology and neurodegeneration in a mouse model (Colgan et al., 2016) and MCI and AD patients’ white matter difference (Fu et al., 2020) while DTI measures did not. In the current study, we found the ODI measure potentially reflect very early microstructural differences associated with the earliest stages of AD pathology. Interestingly, in other studies ODI has been shown to positively correlated with a PET biomarker of postsynaptic density (Mak et al., 2021), thus it is plausible to speculate that the ODI measures in this study are suggestive of very early loss of synaptic density in cognitively intact APOE ε4 carriers. This hypothesis will be the subject of our future investigations in rodent models of AD.

The NODDI model has important limitations that merit mention within the context of our results. For example, gray matter tissue is biologically more heterogenous than white matter as it comprises cell bodies, synapses, dendrites, among other cellular structures, and cells actively exchange substances, including water, for nutrient uptake and waste removal. NODDI is unable to account for the diffusion signal contribution from these constituent microstructures, which may render gray matter too complex for NODDI’s simplified model to accurately quantify the diffusion signal from dendrites alone. The long-term goal of our work is to improve our *in vivo* imaging methods to detect subtle changes in neurite microstructure associated with the pathogenesis of AD. Our future work will focus on the collection of larger datasets that support more sophisticated models of diffusion in brain, such as the Neurite Exchange Imaging (Jelescu et al., 2022) and the Soma and Neurite Density Imaging (Palombo et al., 2020) models, which account for exchange of water between compartments and the diffusion signal contribution from the soma, respectively.

In sum, the present study reports that cognitively intact carriers of the APOE ε4 allele demonstrated lower microstructural integrity in hippocampal subregions that were associated with delayed verbal memory performance. This finding was not evident in the noncarrier group, suggesting that this relationship may represent early pathological changes in the at-risk group. The results of this study align with previous neuroimaging work that report that hippocampal subregions are associated with delayed verbal memory in other populations. Future work is needed to determine the clinical utility of dMRI as a biomarker of the subclinical progression of AD.

## Data Availability

All data produced in the present study may/may not be available upon reasonable request to the authors.

## Acknowledgements

The authors thank Dr. Brian Gordon for generously providing his older adult brain atlas and guidance on coregistration methodology during image preprocessing.

## Supplementary Data

**Figure S1.**
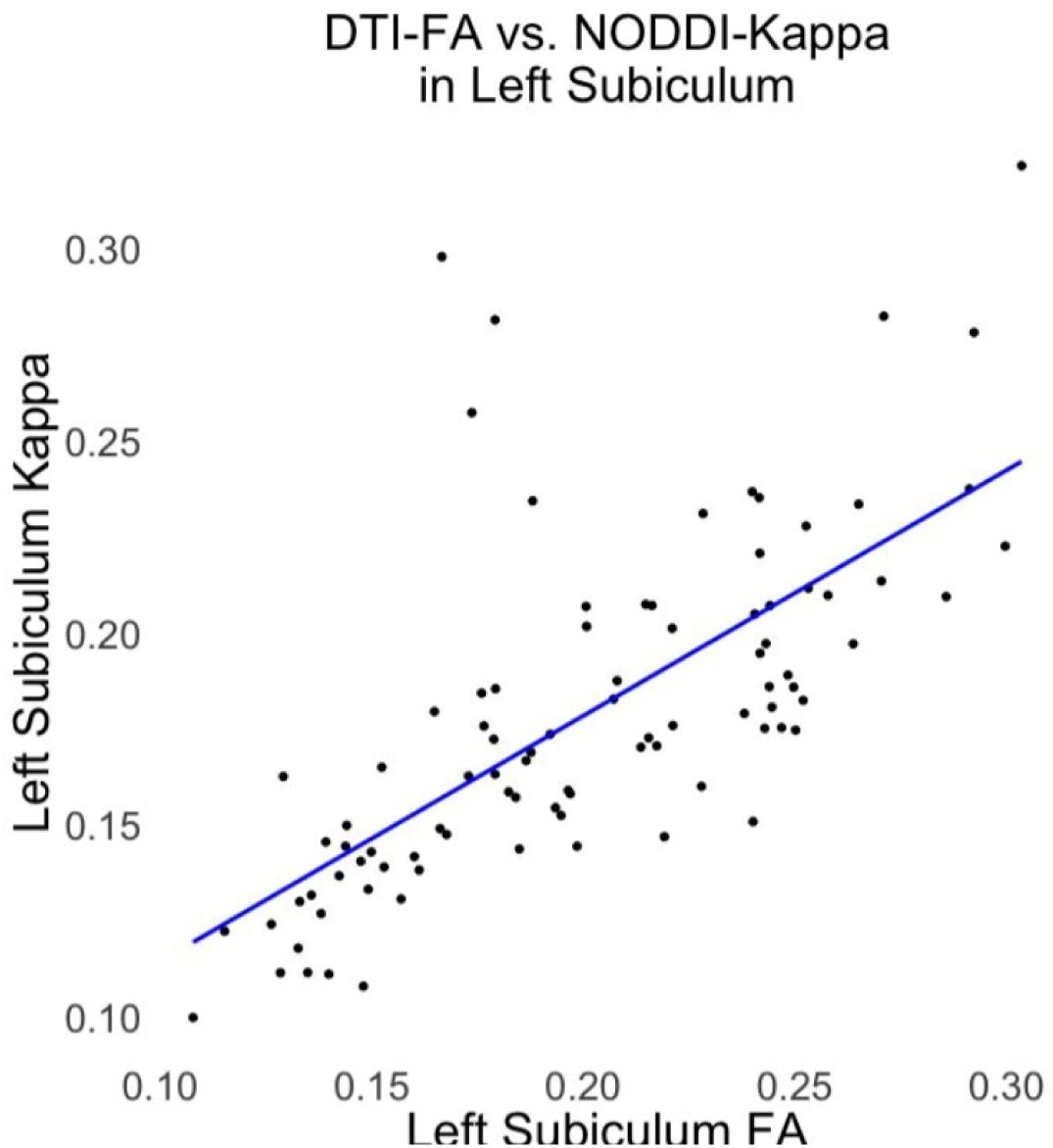
Scatter plot illustrating the relationship between DTI-FA and NODDI-Kappa in the left subiculum.

